# Assessment of Tap Water Quality and Health Risks in Mymensingh City, Bangladesh

**DOI:** 10.1101/2025.10.22.25338587

**Authors:** F.K. Sayema Tanzia, Md. Rased Hasan Sojib, Effat Jahan, Md. Monabbir Hossain, Md. Al-Amin Khan, Mahmudul Hasan, Md. Tariqul Islam, Sabrin Bashar

**Affiliations:** Department of Environmental Science and Engineering, Jatiya Kabi Kazi Nazrul Islam University, Trishal, Mymensingh-2220, Bangladesh; Department of Environmental Science, Bangladesh Agricultural University, Mymensingh-2202, Bangladesh; Faculty of Engineering and Applied Sciences, Cranfield Environmental Centre, Cranfield University, Cranfield, United Kingdom; Department of Pediatrics, University of Alberta, Edmonton, T6G1C9, Alberta, Canada

**Keywords:** Tap water quality, Heavy metal contamination, Health risk assessment, Coliform bacteria, Water supply management

## Abstract

This study assesses the domestic water supply status, physicochemical quality, and associated health risks in a region reliant on groundwater-derived tap water. A cross-sectional survey revealed that 89% of households depend on tap water, yet 78% express significant concerns about its safety, prompting inconsistent treatment practices (45% treat water “seldom”). Physicochemical analysis of tap water samples identified critical contamination issues: 33.3% of samples exceeded Bangladesh (BD ECR 2023) iron (Fe) limits (0.3-2.05 mg/L), while 100% surpassed WHO guidelines (0.3 mg/L). Manganese (Mn) exceeded permissible levels (0.1 mg/L) in 33.3% of samples. Total coliform contamination (up to 12 CFU/100ml) was widespread, though fecal coliforms were absent. Principal Component Analysis (PCA) attributed 82.22% of water quality variance to dissolved solids (electrical conductivity, salinity) and metal contamination (Fe, Mn), linked to geogenic/anthropogenic sources. Health risk assessments revealed non-carcinogenic hazards, particularly for children, with Hazard Index (HI) values reaching 2.87-far exceeding the safety threshold (HI > 1). Iron posed the greatest risk (THQ up to 2.44 for children), underscoring vulnerabilities due to physiological sensitivity. Despite 67% of respondents reporting satisfaction with water quality, stark disparities exist between perception and analytical results, driven by aesthetic/metallic concerns. Urgent interventions are needed, including infrastructure upgrades to curb pipe corrosion, advanced metal removal filtration, and community water treatment education. Policymakers must prioritize stricter industrial regulations and routine monitoring. This study highlights the imperative to align public trust with scientific evidence to safeguard health in groundwater-dependent communities.

## 1. Introduction

Water is a basic resource that is necessary to support life, and contamination of it poses serious risks to public health everywhere. Although having access to safe and clean drinking water is essential, water quality is still a big challenge in many places, including Bangladesh. Many infections spread through contaminated water, causing major health issues, especially in developing countries where access to clean drinking water is sometimes scarce [1], [2]. According to studies, drinking contaminated water causes over 80% of diseases and two-thirds of fatalities in underdeveloped countries, leading to a large loss of productivity and higher healthcare costs [3], [4], [5]. The presence of harmful contaminants, including pathogenic microorganisms, heavy metals, and chemical pollutants, in tap water presents significant health hazards [6], [7]. These contaminants may arise from insufficient water treatment, industrial effluents, agricultural runoff, or deficient sanitation systems. Research indicates that the ingestion of contaminated water is a principal cause of gastrointestinal diseases and waterborne infections such as cholera, typhoid, dysentery, and gastroenteritis [8], [9], [10]. It is estimated that unsafe drinking water accounts for up to 75% of waterborne diseases globally, with the World Health Organization (WHO) attributing nearly 80% of illnesses in developing countries to inadequate sanitation or water contamination [11], [12].

In Bangladesh, concerns over tap water quality have grown as a result of issues such as inadequate water treatment, rising urbanization, and environmental contamination [13]. Poor drinking water quality raises the risk of infectious illnesses, which disproportionately impact poor groups. Previous research has identified microbial pollution, high amounts of heavy metals, and other physicochemical abnormalities in tap water [14], [15]. Furthermore, issues such as inadequate water storage, agricultural practices, and a lack of public knowledge worsen water quality degradation [16].

Given these pressing concerns, this study aims to assess the current status of water supply and evaluate the physicochemical and bacteriological quality of tap water samples collected from various sources in and around Mymensingh City, Bangladesh. By analyzing key water quality parameters and identifying potential sources of contamination, the research seeks to provide valuable insights into the challenges faced by the region’s water supply infrastructure. The findings of this study will contribute to evidence-based recommendations for improving water quality management, ensuring public health safety, and guiding future policy interventions. Ensuring access to safe and potable water is critical for sustaining public health and well-being. This study will bridge knowledge gaps regarding water quality issues in Mymensingh City by conducting a comprehensive assessment of tap water samples. By identifying contamination sources and evaluating water safety against national and international standards, the study aims to support targeted efforts to improve water quality and mitigate the risk of waterborne diseases in the community.

## 2. Materials and Methods

### 2.1 Study Area

Mymensingh City, a major financial and educational hub in north-central Bangladesh, is situated along the Brahmaputra River (24°43’-24°46’N, 90°23’-90°26’ E) and spans 91.315 km² [17]. Its strategic riverine location exposes it to water quality challenges, including contamination from industrial discharge, agricultural runoff, and urban waste, compounded by high population density and anthropogenic activities such as rapid urbanization and industrialization [18]. As a growing urban center, the city’s reliance on tap water for its residents and institutions like Bangladesh Agricultural University underscores the urgency of assessing physicochemical and bacteriological parameters to ensure safe water supply, public health, and sustainable development. This study focuses on Mymensingh due to its socio-economic significance, vulnerability to water quality degradation, and the critical need for improved water management strategies.

**Figure 1:**
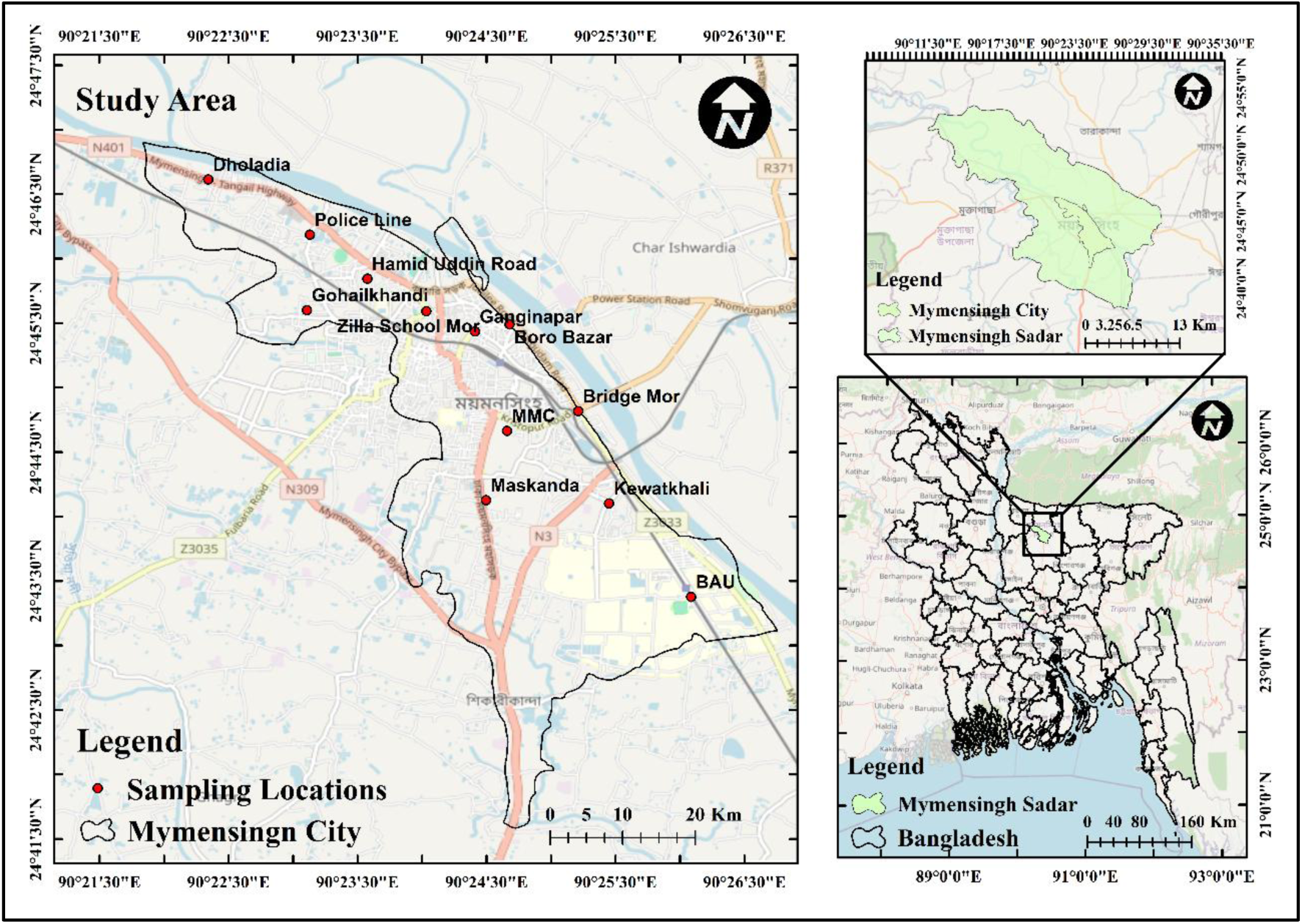
Map of the Study Area.

### 2.2 Sampling locations and sample size

In this study, a total of 12 tap water samples were collected from different locations across Mymensingh City to provide an initial assessment of water quality and associated health risks. While this sample size may appear limited, the sampling points were strategically selected to represent various socio-economic zones, household types, and proximity to key water supply sources (e.g., municipal and private wells). This approach aimed to ensure a degree of spatial representativeness within logistical and resource constraints. However, we acknowledge that a larger sample size could enhance statistical robustness and better capture local heterogeneity.

Samples were collected from 12 areas inside Mymensingh city, including Dholadia, Boro Bazar, Hamid Uddin Road, Mymensingh Medical College (MMC), Ganginapar, Zilla School Mor, Maskanda, Bangladesh Agricultural University (BAU), Bridge Mor, Police Line, Kewatkhali, and Gohailkhandi (**Table 1**).

**Table 1:** Geographical Coordinates of Tap Water Sampling Locations in Mymensingh City.

| Sample ID | Location Name | Latitude | Longitude |
| --- | --- | --- | --- |
| 1 | Dholadia | 24.776888 | 90.372399 |
| 2 | Boro Bazar | 24.758157 | 90.411322 |
| 3 | Hamid Uddin Road | 24.764060 | 90.392963 |
| 4 | Mymensingh Medical College | 24.744403 | 90.410993 |
| 5 | Ganginapar | 24.757250 | 90.406853 |
| 6 | Zilla School Mor | 24.759859 | 90.400567 |
| 7 | Maskanda | 24.735435 | 90.408273 |
| 8 | Bangladesh Agricultural University | 24.722931 | 90.434782 |
| 9 | Mymensingh Bridge Mor | 24.746948 | 90.420187 |
| 10 | Police Line | 24.769756 | 90.385538 |
| 11 | Kewatkhali Power House Road | 24.735033 | 90.424177 |
| 12 | Gohailkhandi | 24.760002 | 90.385149 |

### 2.3 Data Collection

Primary data were collected through surveys and on-site field measurements between January and February 2024. Surveys involved structured questionnaires administered to over 100 participants across selected zones, capturing community insights on water usage and quality concerns. Concurrently, physicochemical parameters- pH, electrical conductivity (EC), total dissolved solids (TDS), and dissolved oxygen (DO) were measured using portable instruments. The HI-98194 Multiparameter Meter was employed for pH, EC, and TDS following calibration with distilled water and buffer solutions (pH 7.0). DO was measured using the HACH HQ30d meter. For each parameter, 50 mL samples were analyzed in triplicate after rinsing probes with distilled water to ensure accuracy [19].

All water samples were collected during the morning hours (between 8:00 AM and 10:00 AM) to maintain consistency in sampling time. Prior to sample collection, taps were flushed for approximately 3-5 minutes to ensure that stagnant water in the pipes was cleared and that the water sampled reflected actual supply conditions. This flushing also helped minimize variability due to water that had been sitting overnight in household plumbing systems.

Moreover, this study was conducted during the dry season, which may not fully reflect seasonal fluctuations in water quality. Previous studies in similar climatic regions have shown that water contamination, particularly microbial and metal concentrations, can vary significantly between the monsoon and dry seasons due to surface runoff, rising water tables, and changes in water treatment efficiency [20], [21]. Therefore, future studies should incorporate longitudinal sampling across different seasons to provide a more comprehensive understanding of temporal dynamics in water quality and associated health risks.

### 2.4 Laboratory Analysis

Water samples were analyzed at the Zonal Laboratory of the Department of Public Health Engineering, Mymensingh. Physicochemical analysis determined iron (Fe) and manganese (Mn) concentrations using an Atomic Absorption Spectrophotometer (AAS) (PerkinElmer, Model XYZ) following APHA Method 3111D [22]. Quality control procedures included the use of calibration curves prepared with at least five standard solutions for each element, with correlation coefficients (R²) above 0.999. Analytical accuracy was verified using certified reference materials (CRMs) from the National Institute of Standards and Technology (NIST). Procedural blanks and triplicate analyses were performed to assess contamination and precision. The method detection limits (MDLs) were 0.01 mg/L for Fe and 0.005 mg/L for Mn. Bacteriological analysis quantified Total Coliform (TC) and Fecal Coliform (FC) via the Membrane Filtration Method (APHA 9222B), with results expressed in colony-forming units per 100 mL (CFU/100 mL) [23].

### 2.5 Data Analysis and Interpretation

Data for this study were collected from both primary and secondary sources and organized in tabular format using Microsoft Office (MS) Word and MS Excel. The analysis was carried out based on the study objectives, incorporating correlation analysis and Principal Component Analysis (PCA) using the statistical software R (version 4.4.2). Inverse Distance Weighting (IDW) was performed for spatial analysis using ArcGIS (version 10.8).

### 2.6 Inverse Distance Weighting (IDW) Interpolation Method

The Inverse Distance Weighting (IDW) interpolation method was applied for spatial analysis. IDW is a probabilistic interpolation technique that uses a weighted linear combination of values from known locations to estimate unknown values [24]. The underlying principle of IDW is that closer points have a greater impact on the estimated value than those farther away. The interpolation is calculated using the following (Equation. 1):

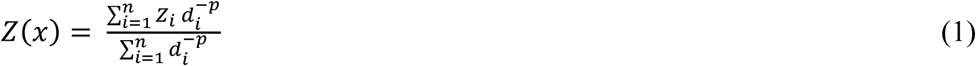

where:

Z(x) is the predicted value at an unknown location; Z_i_ represents the observed values at known points; d_i_ is the distance between the known point *i* and the location *x*; p is the power parameter that controls how strongly distance influences weighting, and n is the number of known points considered in the interpolation.

As shown in (Equation. 1), the weighting factor 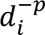 determines how much influence each known point has on the estimated value. When p is higher, the interpolation gives more weight to closer points, producing a more localized effect. Conversely, a lower p value results in a smoother interpolation where distant points have a greater influence. The choice of p is critical, as it directly affects the accuracy of the spatial analysis [25]. IDW is widely used in GIS, environmental science, and geostatistics for spatial interpolation, such as predicting temperature, rainfall, soil properties, and pollution levels [26].

#### 2.6.1 Cross-Validation of IDW Interpolation

Cross-validation was performed to assess the accuracy of the IDW interpolation results. This was done using the Geostatistical Analyst extension in ArcMap 10.8. Each data point was temporarily removed, and its value was predicted using the remaining points. The observed and predicted values were then compared to calculate error metrics such as mean error (ME) and root mean square error (RMSE) [27]. These metrics helped evaluate the model’s performance and identify any systematic bias [28]. Cross-validation was carried out using the Inverse Distance Weighted (IDW) interpolation method in ArcMap 10.8 to assess the accuracy of spatial predictions for Fe and Mn concentrations. Each observed value was removed one at a time, and its value was predicted using the surrounding data points. The predicted values were then compared with the actual observed values to evaluate the performance of the interpolation. The cross-validation results are shown in **Table 2**. The mean observed concentration was 0.96 mg/L, while the mean predicted value was slightly lower at 0.84 mg/L. A mean error (ME) of −0.117 was recorded, which indicates that the model slightly underestimated the actual concentrations. The root mean square error (RMSE) was 0.579. This value suggests that the IDW model provided a moderate level of prediction accuracy for Fe distribution. In the case of Mn (Manganese), the mean observed concentration was 0.396 mg/L, and the mean predicted concentration was 0.314 mg/L. A mean error of −0.082 was found, again pointing to a slight underestimation by the model. The RMSE value for Mn was 0.574, similar to that of Fe, and indicates a comparable level of interpolation accuracy.

**Table 2:** Cross-validation error statistics for Fe (Iron) using IDW interpolation.

| Parameters | Metric | Value | Standard/Benchmark | Interpretation |
| --- | --- | --- | --- | --- |
| **Fe** | Mean Observed | 0.96 |  | Average observed concentration in samples |
|  | Mean Predicted | 0.84 |  | Average value estimated by IDW model |
|  | Mean Error (ME) | - 0.117 | ~ 0 | Indicates bias; close to 0 means unbiased |
|  | RMSE | 0.579 | Lower = Better | Moderate prediction accuracy |
| **Mn** | Mean Observed | 0.396 |  | Average observed concentration in samples |
|  | Mean Predicted | 0.314 |  | Average value estimated by IDW model |
|  | Mean Error (ME) | -0.082 | ~ 0 | Indicates bias; close to 0 means unbiased |
|  | RMSE | 0.574 | Lower = Better | Moderate prediction accuracy |

The results suggest that the IDW method performed reasonably well for both Fe and Mn. Although slight underestimation was observed in both cases, the errors remained within an acceptable range for environmental spatial analysis. These findings support the use of IDW as a suitable method for estimating the spatial variation of heavy metal concentrations in the study area.

### 2.6 Health Risk Assessment

This study focuses on chronic risk assessment, with the health risk assessment (HRA) of heavy metals or metalloids primarily based on evaluating the intensity of potential risks. Only Fe and Mn were considered for health risk assessment, as other potentially toxic elements (e.g., Pb, As, Cr) were not analyzed due to analytical and resource constraints. Future studies should include a broader range of elements for a more comprehensive assessment. These risks are expressed in non-carcinogenic or carcinogenic health effects [29]. The key toxicity factors considered include the reference dose (RfD) used to characterize non-carcinogenic risks [30]. The process began by selecting iron (Fe) and Manganese (Mn) to evaluate their potential health risks on specific population groups, including children, adult females, and adult males. These groups were differentiated based on various characteristics, such as exposure frequency, exposure duration, ingestion rate, and body weight [31].

The exposure assessment included calculating the average daily intake (ADI), expressed in mg kg^−1^day^−1^, for ingestion. This was determined using the formula equation (2):

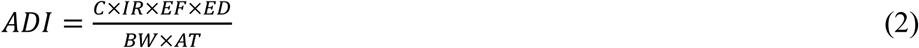

The explanation of the parameters and the values used in the calculations are provided in Table 1. The hazard quotient (HQ), which evaluates risk for ingestion exposure, is calculated as follows equation (3):

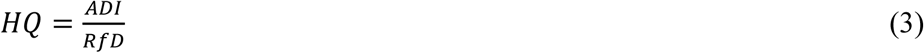

The explanation and values of the RfD for Fe and Mn are presented in Table 2. The total hazard index (HI), which assesses the overall non-carcinogenic risk, is calculated using the following equation (4):

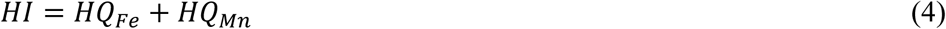

**Table 3.**
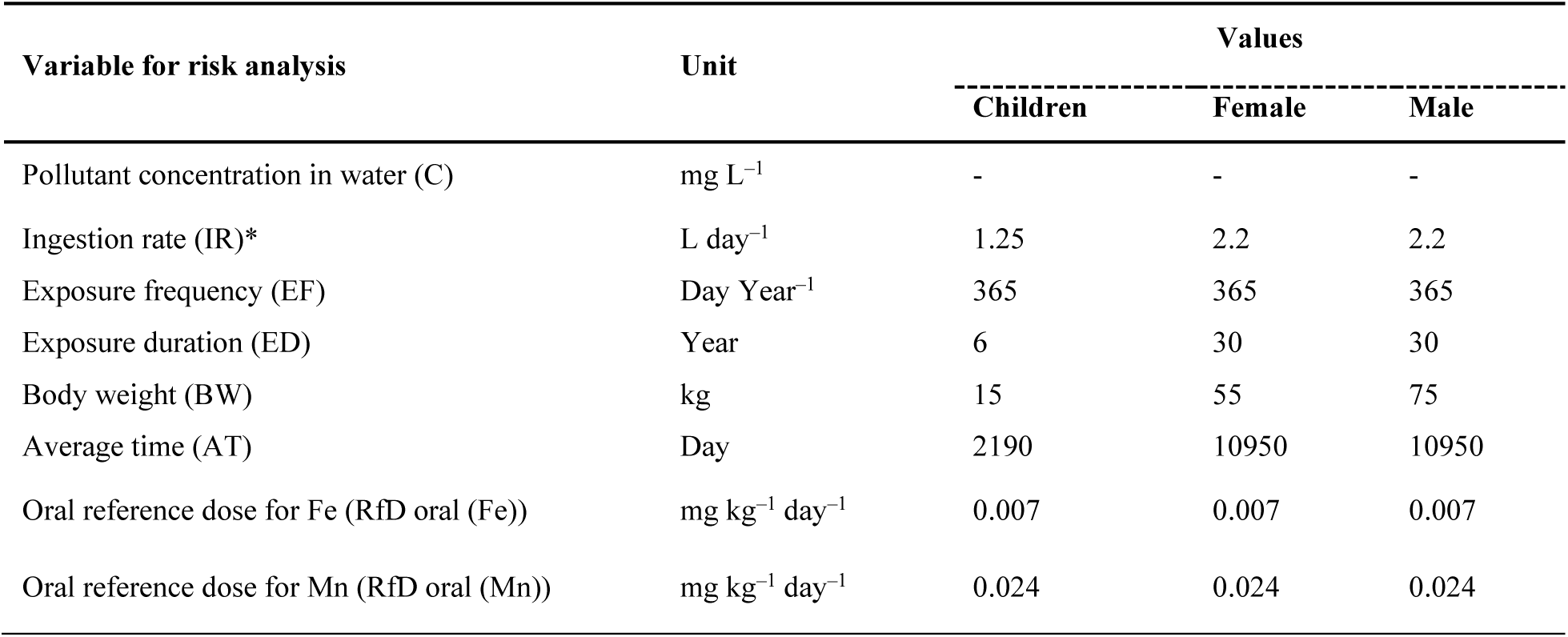
Summary of variable used to calculate Human Health Risk Assessment [29].

The health risk assessment was based on deterministic (point estimate) methods using mean concentrations of Fe and Mn. Although this approach provides a general indication of risk, it does not capture the variability and uncertainty inherent in environmental exposure data. The relatively high standard deviations observed (**Table 4**) suggest significant spatial or temporal variability. A probabilistic approach, such as Monte Carlo simulation, would offer a more robust evaluation by incorporating the range and distribution of input variables. Due to resource and computational limitations, such analysis was not included in the current study. We recommend that future studies adopt probabilistic risk assessment methods to better quantify uncertainty and variability in exposure and risk estimates.

## 3. Results and Discussions

### 3.1 Present water supply status

#### 3.1.1 Primary Source of Water for Domestic Use

The study found that tap water is the major home water supply for 89% of respondents, while hand tubewells service the remaining 11% (Figure 2). Tap water is normally supplied from subsurface aquifers, stored in above tanks, and transported via pipes, whilst hand tubewells directly access groundwater using manual pumping methods. These aquifers undergo extraction, storage in overhead tanks, and subsequent distribution through municipal pipelines, ensuring a relatively continuous and controlled water supply (WHO, 2017). Households that rely on hand tubewells extract groundwater manually using simple pump mechanisms. While this approach provides an independent and cost-effective water source, it is often more vulnerable to contamination and fluctuations in groundwater levels, particularly during dry seasons [33]. The low percentage of respondents using hand tubewells suggests a shift towards municipal water systems, likely due to better infrastructure, concerns over groundwater quality, and decreasing groundwater availability [34].

**Figure 2:**
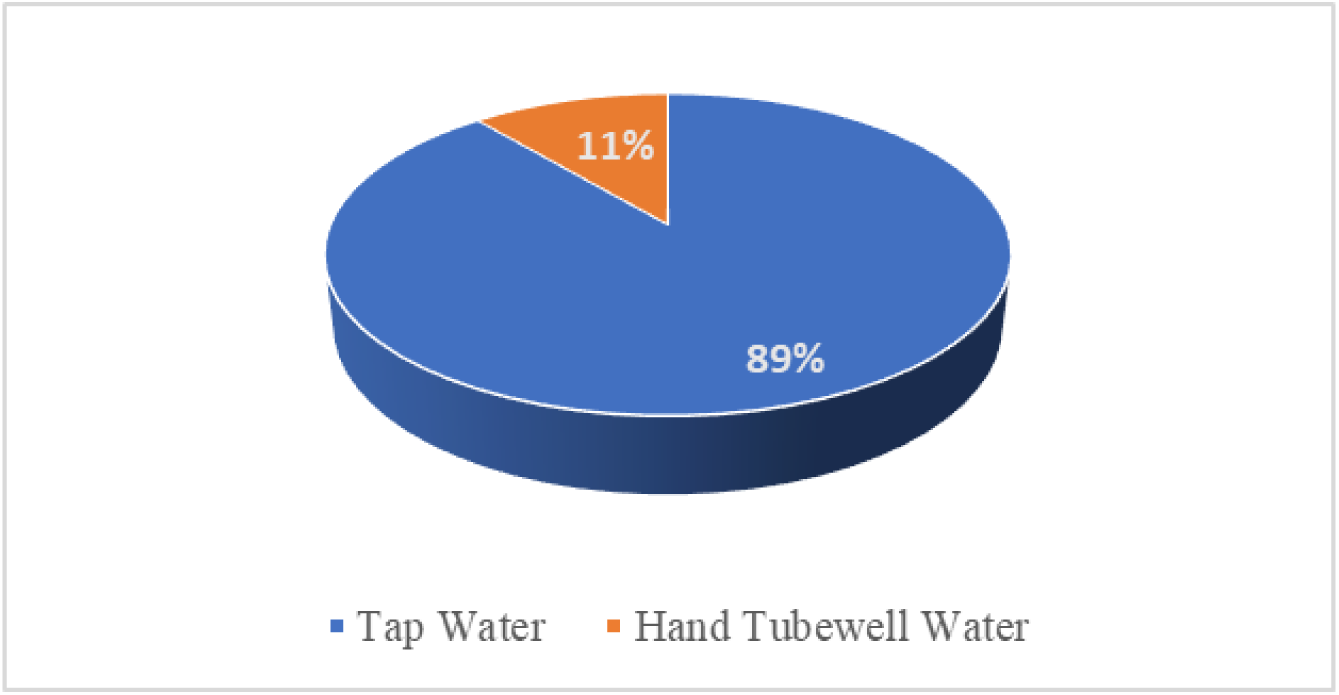
Primary Source of Water for Domestic Use (%)

#### 3.1.2 Tap Water Usage and Drinking Habits

The data shows that tap water usage varies among respondents, with 22% using it for more than four hours a day, and 56% using it for one to two hours, mainly for cooking, cleaning, and hygiene (Mekonnen & Hoekstra, 2016). Larger households or those with jobs requiring significant water use may account for the higher usage beyond four hours [35]. When it comes to drinking habits, 78% of respondents rely on tap water, while 11% never use it, and another 11% use it infrequently (Figure 3b). This indicates a general trust in the water supply system, though concerns about water quality and infrastructure persist [36], [33]. To ensure public health, it is essential to regularly test water quality and continue improving water treatment systems [32].

**Figure 3:**
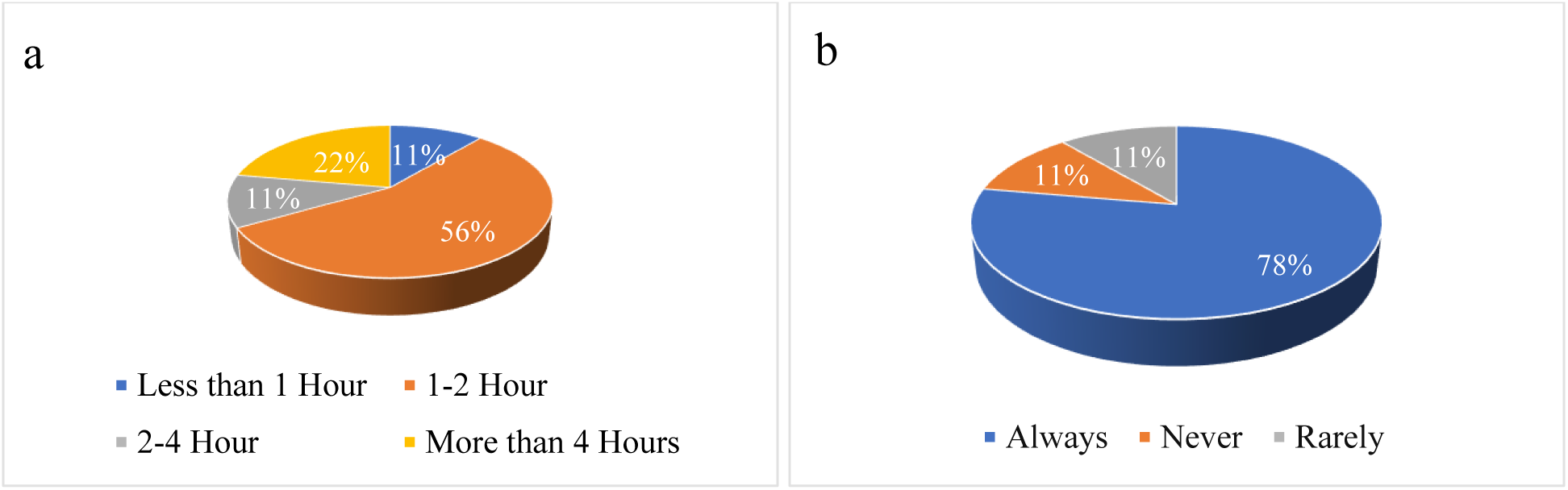
(a) Tap water usage in a day (%); (b) Tap water for drinking purposes (%)

#### 3.1.3 Perception and Satisfaction with Tap Water Quality

The evaluation of water quality satisfaction reveals that 67% of respondents expressed satisfaction, while 11% remained neutral, and 22% reported dissatisfaction. This distribution suggests that a significant portion of the population is content with the water quality. The high percentage of satisfied respondents highlights the effectiveness of efforts to ensure access to clean and safe water, fostering trust and confidence among the community (Figure 4a). However, in a recent poll on tap water purification before consumption, a considerable 78% of respondents indicated being extremely worried about the problem, while a smaller 11% remained indifferent or slightly concerned. This heightened worry likely arises from greater knowledge of water quality and safety, impacted by variables such as environmental pollution, reporting of pollutants in water sources, and increased public consciousness of health hazards connected with untreated water (Figure 4b). The increasing concern could be attributed to several factors, including rising awareness of environmental pollution, frequent media reports on contamination incidents, and a broader societal shift toward prioritizing health and water quality [36].

**Figure 4:**
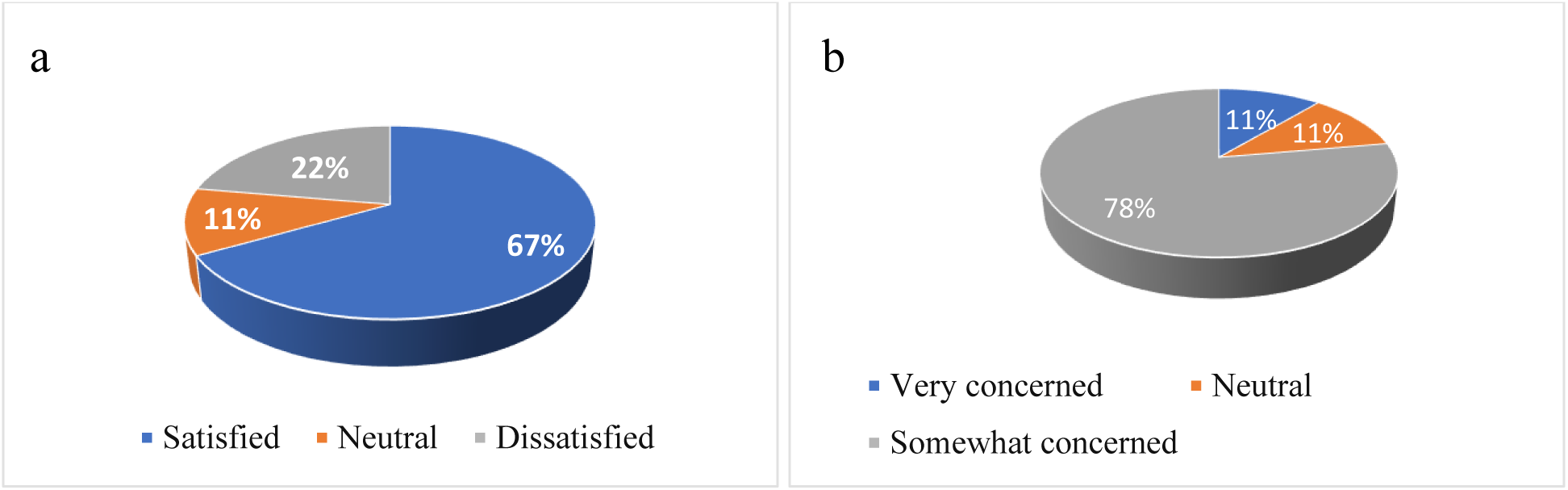
(a) Satisfaction of Water Quality (%); (b) Concerned about tap water quality (%)

#### 3.1.4 Tap Water Treatment and Perceived Changes

Survey participants exhibit varying behaviors with respect to their current tap water treatment practices prior to consumption. For example, 11% of them do not treat their water at all, 22% treat it occasionally, 45% treat it seldom, and 22% treat it consistently. Inadequate knowledge of appropriate treatment techniques may be the cause of the uneven adoption of treatment procedures against pollutants (Figure 5a). Interestingly, these treatment habits align with perceptions of water quality over time. According to the survey, only 56% of respondents believe that the purity of municipal water has deteriorated, while 33% have maintained their stance and 11% are uncertain (Figure 5b). The prevalent view of declining water quality among respondents may be ascribed to growing pollution, aged infrastructure leading to pipe corrosion, and heightened awareness of water safety problems due to reported occurrences. These issues combined add to increased concerns surrounding tap water dependability and safety [33].

**Figure 5:**
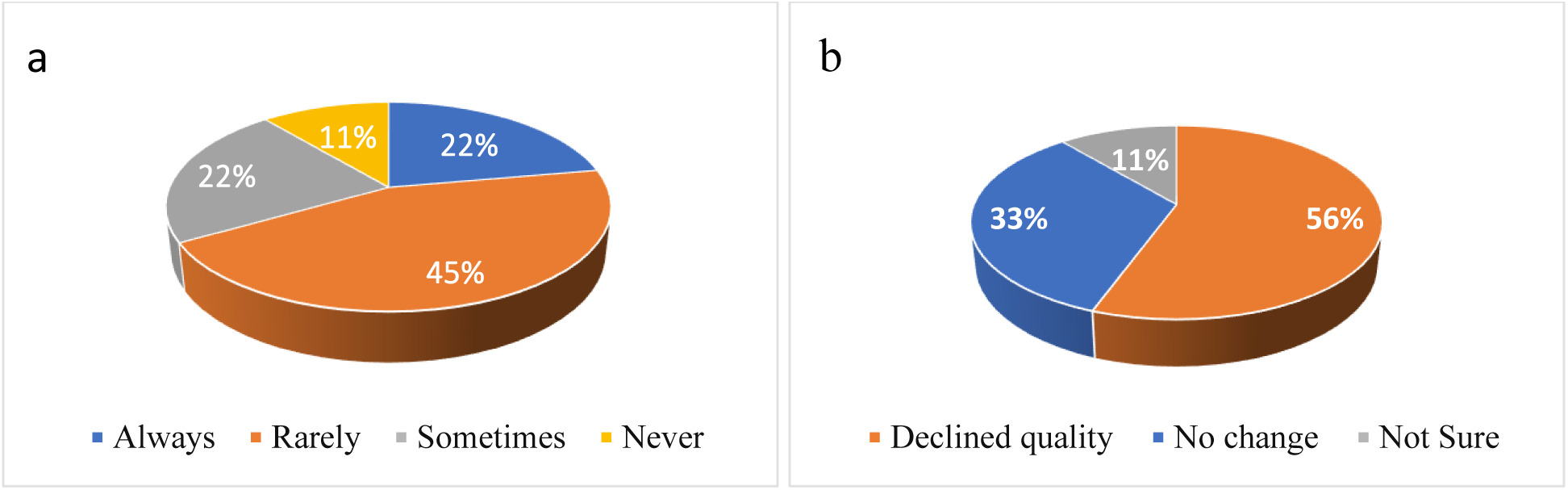
(a) Tap water treatment before consumption (%); (b) Changes in the tap water quality over the past few years (%)

### 3.2 Physicochemical parameters of tap water sample

A comprehensive assessment of water quality parameters was conducted, and the summary statistics are presented in **Table 4**. The results indicate variations in physicochemical characteristics, with some parameters exceeding national and international permissible limits.

The relatively high standard deviations for Fe (±0.52 mg/L) and Mn (±0.51 mg/L) across 12 samples suggest substantial variability in tap water quality across different locations. This spatial heterogeneity reflects uneven distribution of contaminants possibly influenced by varying pipe materials, water sources, or maintenance issues. While the analytical procedures ensured internal consistency (as described in Section 2.4), the observed variation is more likely due to genuine environmental variability rather than measurement imprecision. Nonetheless, the small sample size (n = 12) may limit the statistical power, and results should be interpreted with caution.

**Table 4:** Summary Statistics of Water Quality Parameters and Compliance Assessment.

| Parameter | Range | Mean ± SD | Skewness | Kurtosis | Standard value | | % Exceeding the limits | |
| --- | --- | --- | --- | --- | --- | --- | --- | --- |
|  |  |  |  |  | BD (ECR 2023) standard | WHO (2011) standard | BD (ECR 2023) standard | WHO (2011) standard |
| pH | 6.95 - 7.52 | 7.26 ± 0.16 | -0.239 | -0.012 | 6.5-8.5 | 6.5-8.5 | - | - |
| EC (μS/cm) | 253.00 - 350.00 | 304.17 ± 29.48 | -0.416 | -0.557 | 2000 | 750 | - | - |
| TDS (mg/L) | 126.00 - 175.00 | 152.08 ± 14.85 | -0.456 | -0.495 | 1000 | 500 | - | - |
| Salinity (PSU) | 0.12 - 0.17 | 0.15 ± 0.02 | -0.478 | -0.136 | 0.6 | - | - | - |
| DO (mg/L) | 4.29 - 9.52 | 6.70 ± 1.35 | 0.346 | 1.031 | 6.5-8 | 6.5-8 | 16.7 | 16.7 |
| Fe (mg/L) | 0.55 - 2.05 | 0.96 ± 0.52 | 1.306 | 0.390 | 0.3-1.0 | 0.3 | 33.3 | 100.0 |
| Mn (mg/L) | 0.04 - 1.24 | 0.40 ± 0.51 | 0.874 | -1.397 | 0.1 | 0.1 | 33.3 | 33.3 |

#### 3.2.1 pH

The pH of the water samples ranged from 6.95 to 7.52, with a mean value of 7.26 ± 0.16. This falls within the permissible range of both the Bangladesh Environmental Conservation Rules (ECR, 2023) standard (6.5–8.5) and the World Health Organization (WHO, 2011) guideline (6.5–8.5). The skewness (−0.239) and kurtosis (−0.012) suggest a near-normal distribution of pH values. Since pH influences the solubility of minerals and heavy metals in water, maintaining an optimal range is crucial for aquatic life and human consumption [37].

#### 3.2.2 Electrical Conductivity (EC) and Total Dissolved Solids (TDS)

The EC values ranged from 253 to 350 µS/cm, with a mean of 304.17 ± 29.48 µS/cm, well below the BD (ECR, 2023) standard (2000 µS/cm) and WHO guideline (750 µS/cm). Similarly, TDS values varied from 126 to 175 mg/L, with a mean of 152.08 ± 14.85 mg/L, remaining within acceptable limits (1000 mg/L for BD and 500 mg/L for WHO). These values indicate that the water is fresh and unlikely to pose risks related to high salinity or dissolved solids [38].

#### 3.2.3 Salinity

Salinity values ranged from 0.12 to 0.17 PSU, with a mean of 0.15 ± 0.02 PSU. Since the BD (ECR, 2023) standard limit is 0.6 PSU, all samples remained well within the safe range. The low salinity levels confirm minimal intrusion of saline water, which is essential for maintaining freshwater quality and ecosystem stability [39].

### Dissolved Oxygen (DO)

DO values ranged from 4.29 to 9.52 mg/L, with an average of 6.70 ± 1.35 mg/L. While the majority of samples were within the BD (ECR, 2023) and WHO (2011) standard range (6.5–8.0 mg/L), 16.7% of the samples exceeded the upper limit, indicating potential over-aeration. Higher DO levels can be beneficial for aquatic organisms, but fluctuations might suggest organic pollution or algal activity [40]. Lower DO levels (<6.5 mg/L) may signal organic matter decomposition, which could lead to hypoxic conditions.

#### 3.2.4 Iron (Fe) Contamination

Iron concentrations varied significantly, ranging from 0.55 to 2.05 mg/L, with a mean of 0.96 ± 0.52 mg/L. 33.3% of the samples exceeded the BD (ECR, 2023) standard limit (1.0 mg/L), while 100% exceeded the WHO (2011) permissible limit (0.3 mg/L). High iron levels are a common concern in groundwater, often resulting from the dissolution of iron-bearing minerals or anthropogenic sources such as industrial discharge and agricultural runoff [41]. Elevated iron levels can cause aesthetic issues like staining and unpleasant taste, while long-term exposure may lead to adverse health effects, including gastrointestinal distress and oxidative stress [42]. Excessive iron concentrations exceeding 0.3 mg/L may lead to gastrointestinal distress, including nausea and vomiting, and contribute to conditions such as diabetes and hemochromatosis- a disorder caused by iron overload [43].

#### 3.2.5 Manganese (Mn) Contamination

Manganese levels ranged from 0.04 to 1.24 mg/L, with a mean of 0.40 ± 0.51 mg/L. 33.3% of the samples exceeded both the BD (ECR, 2023) and WHO (2011) standard limit of 0.1 mg/L. High manganese levels in drinking water can pose serious health risks, particularly neurotoxic effects with prolonged exposure [44]. The presence of elevated Mn concentrations could be attributed to natural geological sources or contamination from industrial and agricultural activities [45]. Manganese is an essential element required for the proper functioning of human, animal, and plant metabolism, as it plays a critical role in activating numerous enzymatic processes[46]. Landfills and waste dumps can also contribute to manganese contamination, as decomposing waste leaches Mn into the subsurface [47]. pH and redox potential play a crucial role in Mn mobility, as it becomes more soluble under acidic and reducing conditions, further influencing its release and transport in groundwater systems [48].

#### 3.2.6 Total Coliform (TC)

Total coliform bacteria are indicators of potential contamination in tap water. Comparing the observed TC levels with the Bangladesh standard of 0 CFU/100ml, it’s evident that contamination exists in all sampled locations, as all readings are above the standard. Notably, the highest TC count is recorded at 12 CFU/100ml in Mymensingh Medical College (M-04), followed by 8 CFU/100ml at Mymensingh Bridge Mor (M-09) and 5 CFU/100ml in both Police Line (M-10) and Gohailkhandi (M-12). Conversely, the lowest TC count of 0 CFU/100ml is found at Boro Bazar (M-02). Such elevated levels of total coliform indicate potential health risks, including gastrointestinal illnesses, if consumed without proper treatment [49].

#### 3.2.7 Fecal Coliform

Fecal coliform bacteria, originating from human or animal waste, indicate potential contamination. The Bangladesh standard mandates 0 CPU/100ml for safe drinking water. Fortunately, all tested locations-Boro Bazar, Mymensingh Medical College, Zilla School Mor, Mymensingh Bridge Mor, Police Line, and Gohailkhandi-recorded 0 CPU/100ml, meeting safety standards and confirming no detectable contamination.

#### 3.2.8 The Correlation Analysis of Water Quality Parameters

Correlation analysis is a powerful tool in pollution research, helping to decode the complex nature of human-induced pollution and trace its origins [45]. As highlighted by Attia and Ghrefat (2013), identifying strong correlations among heavy metals within a sample can provide crucial insights into common sources of contamination. The Pearson’s correlation matrix presented in Figure 6 illustrates the relationships among key water quality parameters. The color gradient represents the correlation coefficient (r), ranging from −1.0 (strong negative correlation) to 1.0 (strong positive correlation), with statistical significance levels denoted by asterisks (* p < 0.05, ** p < 0.01, *** p < 0.001).

**Figure 6:**
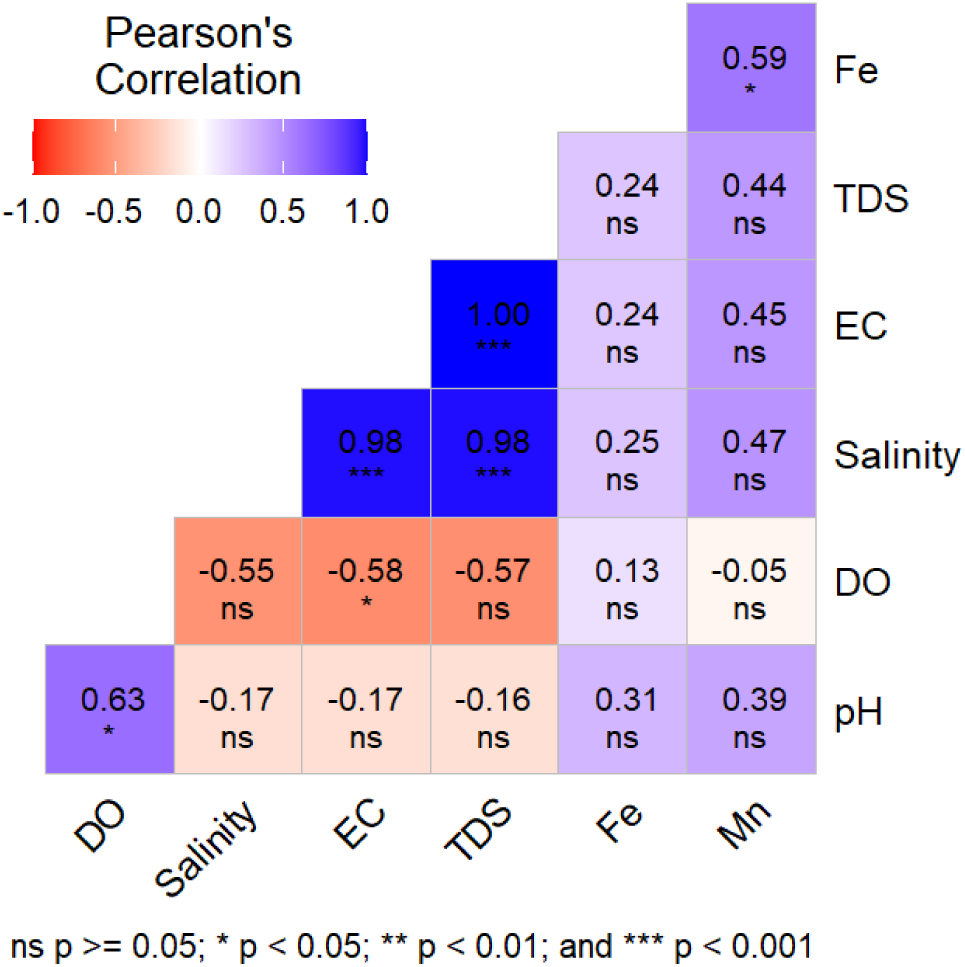
Heat Map Representing Correlation Among Water Quality Parameters.

A strong positive correlation is observed between electrical conductivity (EC) and total dissolved solids (TDS) (r = 0.98, p < 0.001), indicating that increased ion concentration in the water is directly linked to higher TDS values. Similarly, EC also exhibits a strong positive correlation with salinity (r = 0.98, p < 0.001), suggesting that higher conductivity is associated with increased salinity [51]. A statistically significant positive correlation is also observed between dissolved oxygen (DO) and pH (r = 0.63, p < 0.05), implying that oxygen-rich water may tend to have a higher pH. However, Fe exhibits weak correlations with TDS (r = 0.24, p > 0.05) and EC (r = 0.24, p > 0.05), suggesting that its concentration may be influenced by other environmental factors. Likewise, salinity, EC, and TDS show weak negative correlations with DO (−0.55 to −0.58), though only EC shows a statistically significant relationship (p < 0.05), indicating that increased ionic concentration may contribute to reduced oxygen availability.

The observed moderate positive correlation between iron (Fe) and manganese (Mn) concentrations (r = 0.59) suggests that these elements may share common sources or geochemical pathways [44]. In the context of Mymensingh City, this relationship can be attributed primarily to natural geological factors. The region is situated on the alluvial floodplains of the Brahmaputra-Jamuna river system, where groundwater is often rich in iron and manganese due to the weathering of iron- bearing and manganese-rich minerals in the sediment [52], [53], [54]. These elements are commonly mobilized under reducing (anaerobic) conditions in groundwater, which are prevalent in low-lying floodplain areas [55], [56].

Additionally, land use patterns in certain parts of the city, particularly in peri-urban zones, include small-scale metal workshops, vehicle repair yards, and agricultural runoff, all of which may contribute to localized increases in Fe and Mn concentrations [57], [58]. However, without specific industrial discharge data or hydrogeological mapping, the extent of anthropogenic influence remains uncertain. Future research should incorporate geochemical surveys, land use classification, and hydrogeological profiling to better distinguish between natural and anthropogenic sources of these metals. The integration of GIS-based land use maps could also enhance understanding of spatial contamination patterns in relation to industrial clusters or agricultural zones.

#### 3.2.8 Hierarchical Clustering Analysis of Water Quality Parameters

The hierarchical clustering analysis (HCA), as illustrated in Figure 7, enables interpretation of potential contamination sources through cluster patterns among both water quality parameters and sampling locations. In the parameter dendrogram (Figure 7a), two distinct clusters emerge. The first cluster groups salinity, EC, and TDS, suggesting a common geogenic origin likely from the dissolution of subsurface minerals or saltwater intrusion in groundwater [59]. The second cluster, which includes Fe, Mn, DO, and pH, implies a different set of processes dominated by redox-sensitive geochemical reactions, where low-oxygen conditions in aging or corroded pipelines can mobilize Fe and Mn into the water [60]. The clustering of Fe and Mn also supports their moderate correlation (r = 0.59), reinforcing the likelihood of a shared geochemical mobilization pathway, possibly influenced by naturally reducing aquifer environments or bio-corrosion of pipes [61]. On the other hand, the dendrogram of sampling sites (Figure 7b) shows three major groupings, where samples 1, 3, and 6 form a distinct cluster associated with higher metal concentrations suggesting localized contamination hotspots, possibly due to pipeline degradation or suboptimal municipal treatment.

**Figure 7:**
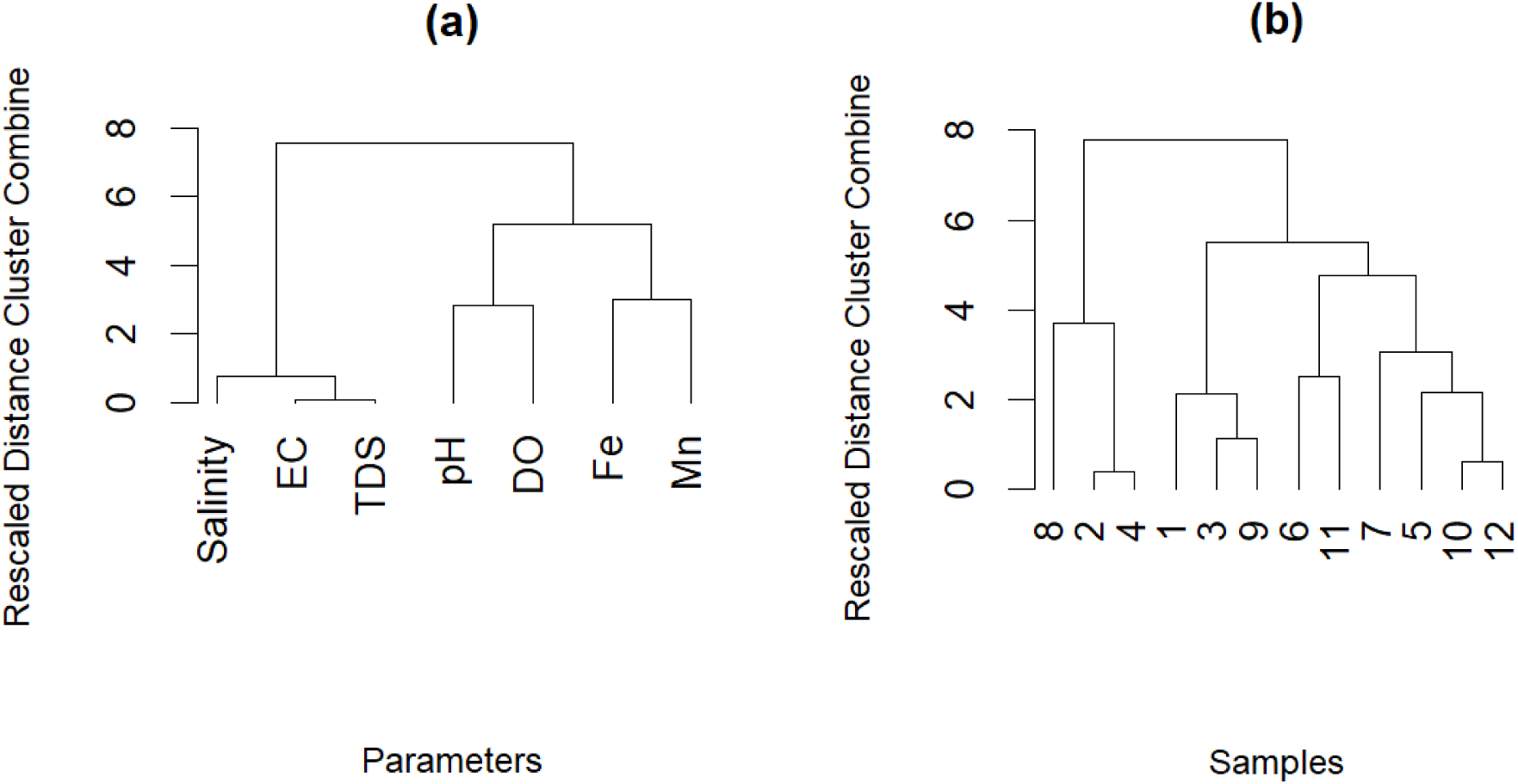
Dendrograms Representing Hierarchical Clustering of (a) Water Quality Parameters and (b) Sample Data.

**Table 4:** Principal Component Analysis (PCA) Results for Water Quality Parameters.

| Parameter | PCA 1 | PCA 2 | PCA 3 | PCA 4 | PCA 5 |
| --- | --- | --- | --- | --- | --- |
| pH | -0.218 | 0.839 | 0.413 | -0.074 | -0.268 |
| EC ($\mu\text{S}/\text{cm}$) | 0.978 | -0.019 | 0.168 | 0.106 | 0.004 |
| TDS ($\text{mg}/\text{L}$) | 0.976 | -0.012 | 0.173 | 0.118 | 0.001 |
| Salinity (PSU) | 0.972 | 0.002 | 0.154 | 0.112 | 0.059 |
| DO ($\text{mg}/\text{L}$) | -0.658 | 0.586 | 0.229 | 0.296 | 0.288 |
| Fe ($\text{mg}/\text{L}$) | 0.322 | 0.717 | -0.548 | 0.267 | -0.105 |
| Mn ($\text{mg}/\text{L}$) | 0.536 | 0.684 | -0.122 | -0.438 | 0.192 |
| Eigenvalues | 3.725 | 2.031 | 0.620 | 0.395 | 0.206 |
| % of Variance | 53.211 | 29.010 | 8.853 | 5.638 | 2.946 |
| Cumulative % of variance | 53.211 | 82.221 | 91.074 | 96.712 | 99.658 |

Extraction Method: Principal Component Analysis.

#### 3.2.9 Principal Component Analysis (PCA) of Water Quality Parameters

Principal Component Analysis (PCA) was performed to identify the major factors influencing tap water quality in Mymensingh City (**Table 4**). The first two principal components (PC1 and PC2) accounted for a cumulative variance of 82.22%, indicating they capture the dominant patterns in the dataset. PC1 (53.21%) had high positive loadings for electrical conductivity (EC = 0.978), total dissolved solids (TDS = 0.976), and salinity (0.972), suggesting that this component primarily represents mineral content and ionic strength, likely associated with natural geogenic processes such as mineral dissolution from alluvial sediments. PC2 (29.01%) showed strong positive loadings for pH (0.839), Fe (0.717), Mn (0.684), and DO (0.586), pointing toward a redox- influenced geochemical signature that may reflect groundwater-metal interactions and possibly the influence of oxidizing or reducing conditions in the distribution system [62], [63], [64].

The remaining components, PC3 (8.85%), PC4 (5.64%), and PC5 (2.95%), together contributed only 17.78% of the total variance and were thus not the primary focus of interpretation. Nonetheless, PC3 showed moderate negative loading for Fe (−0.548) and positive loading for pH (0.413), possibly indicating some localized physicochemical shifts or pipe interactions that alter iron solubility [65]. The lesser components may also reflect secondary or sporadic influences, such as microbial activity, biofilm development, or unmeasured organic pollutants [66]. Although no specific parameters related to organic or anthropogenic pollutants were included in this PCA, future studies should broaden the scope to include organic contaminants, pesticides, and endocrine-disrupting compounds, especially considering their relevance in urbanized environments. Furthermore, the inclusion of geospatial analysis or land-use mapping could help clarify if these weaker components relate to localized pollution events or transient contamination from urban runoff or agricultural areas.

Integrating results from PCA, correlation, and HCA, three primary contamination sources are inferred: (1) geogenic mineral dissolution and ionic enrichment (TDS, salinity, EC), (2) redox-driven mobilization of trace metals (Fe, Mn, DO, pH), and (3) infrastructure-related influences specific to clusters of samples. This multivariate approach enhances source attribution and can inform targeted interventions for improving urban tap water quality.

#### 3.2.10 Spatial Distributions of Iron (Fe) and Manganese (Mn)

The left map (Figure 8(a)) illustrates the spatial distribution of Fe concentrations. The highest Fe levels (red zones) are observed in the Dholadia and Maskanda regions, indicating localized enrichment. The concentration gradually decreases towards the southeastern and central parts of the study area, where lower values (green zones) are recorded. The spatial variation suggests that Fe contamination could be influenced by natural geochemical processes, sediment composition, and potential anthropogenic sources such as industrial and agricultural runoff.

**Figure 8:**
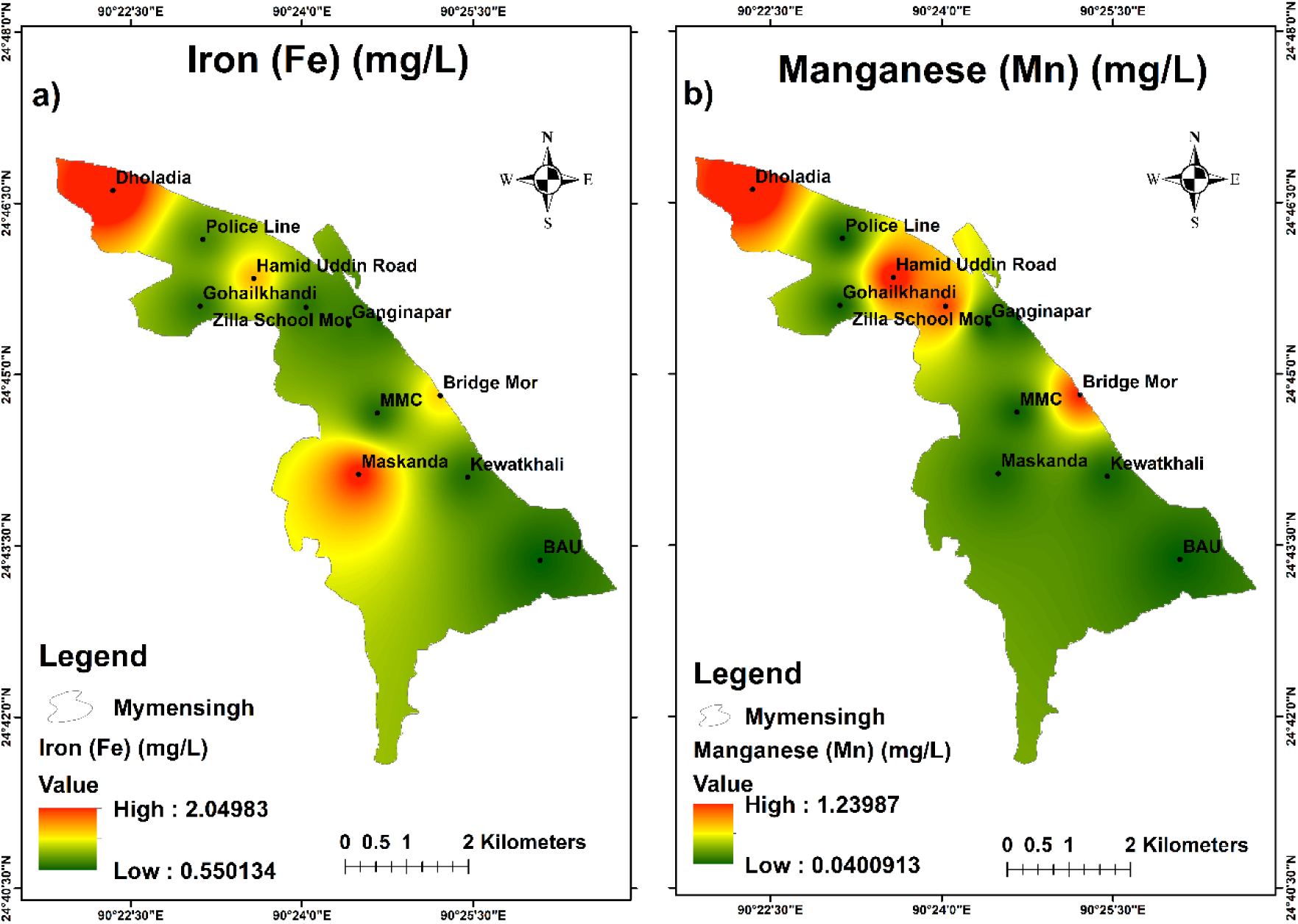
Spatial distributions of the study area (a) Iron-Fe and (b) Manganese-Mn.

The right map (Figure 8 **(b)**) represents the spatial distribution of Mn concentrations. Similar to Fe, Mn exhibits elevated levels in the Dholadia region, with additional hotspots observed near Hamid Uddin Road and MMC. The southeastern region, particularly around BAU, shows relatively lower Mn concentrations. The observed Mn distribution pattern may be attributed to lithological factors, redox conditions, and anthropogenic inputs, including industrial discharge and landfill leachates.

#### 3.2.11 Non-Carcinogenic Health Risk Assessment

The non-carcinogenic health risks associated with Fe and Mn exposure were evaluated using the hazard quotient (THQ) and hazard index (HI), with results presented in **Table 5**.

**Table 5:** Chemical Species-Wise Hazard Quotient and Non-Cancerous Risk Assessment.

| Sample ID | Chemical species-wise total hazard quotient (THQ) |  |  |  |  |  | Non-cancerous risk |  |  |
| --- | --- | --- | --- | --- | --- | --- | --- | --- | --- |
|  | THQ children |  | THQ adult female |  | THQ adult male |  | Hazard Index (HI) |  |  |
|  | Fe | Mn | Fe | Mn | Fe | Mn | Children | Female | Male |
| 1 | 2.44E+01 | 4.31E+00 | 1.17E+01 | 2.07E+00 | 8.59E+00 | 1.52E+00 | 2.87E+01 | 1.38E+01 | 1.01E+01 |
| 2 | 7.38E+00 | 1.39E-01 | 3.54E+00 | 6.67E-02 | 2.60E+00 | 4.89E-02 | 7.52E+00 | 3.61E+00 | 2.65E+00 |
| 3 | 1.59E+01 | 3.89E+00 | 7.63E+00 | 1.87E+00 | 5.59E+00 | 1.37E+00 | 1.98E+01 | 9.50E+00 | 6.96E+00 |
| 4 | 7.26E+00 | 1.74E-01 | 3.49E+00 | 8.33E-02 | 2.56E+00 | 6.11E-02 | 7.44E+00 | 3.57E+00 | 2.62E+00 |
| 5 | 7.98E+00 | 2.08E-01 | 3.83E+00 | 1.00E-01 | 2.81E+00 | 7.33E-02 | 8.18E+00 | 3.93E+00 | 2.88E+00 |
| 6 | 7.62E+00 | 3.30E+00 | 3.66E+00 | 1.58E+00 | 2.68E+00 | 1.16E+00 | 1.09E+01 | 5.24E+00 | 3.84E+00 |
| 7 | 2.12E+01 | 2.78E-01 | 1.02E+01 | 1.33E-01 | 7.46E+00 | 9.78E-02 | 2.15E+01 | 1.03E+01 | 7.56E+00 |
| 8 | 6.55E+00 | 1.39E-01 | 3.14E+00 | 6.67E-02 | 2.30E+00 | 4.89E-02 | 6.69E+00 | 3.21E+00 | 2.35E+00 |
| 9 | 1.48E+01 | 3.51E+00 | 7.09E+00 | 1.68E+00 | 5.20E+00 | 1.23E+00 | 1.83E+01 | 8.77E+00 | 6.43E+00 |
| 10 | 8.57E+00 | 1.74E-01 | 4.11E+00 | 8.33E-02 | 3.02E+00 | 6.11E-02 | 8.75E+00 | 4.20E+00 | 3.08E+00 |
| 11 | 7.38E+00 | 2.08E-01 | 3.54E+00 | 1.00E-01 | 2.60E+00 | 7.33E-02 | 7.59E+00 | 3.64E+00 | 2.67E+00 |
| 12 | 7.62E+00 | 1.91E-01 | 3.66E+00 | 9.17E-02 | 2.68E+00 | 6.72E-02 | 7.81E+00 | 3.75E+00 | 2.75E+00 |

The analysis of non-cancerous health risks associated with iron (Fe) and manganese (Mn) exposure across 12 samples reveals significant concerns, particularly for children. The Total Hazard Quotient (THQ) and Hazard Index (HI) metrics, which quantify risk levels (values >1 indicate potential harm), show that children face disproportionately high risks. For all samples, HI values for children range from 6.69 to 28.7, with Fe contributing 79-95% of the total risk. Sample 1 stands out as the most hazardous, with a child HI of 28.7 driven by an exceptionally high Fe THQ of 24.4. While Mn poses a secondary risk overall, it becomes notable in specific samples (e.g., Samples 3, 6, and 9), where child Mn THQ values reach 3.89-3.30. Adults also face elevated risks, though less severe than children. Adult females exhibit HI values of 3.21-13.8, while males range from 2.35-10.1, with Fe accounting for 70-90% of the risk in both groups.

The calculated Hazard Index (HI) for children, reaching up to 28.7, indicates a severe potential for non-carcinogenic health effects, particularly due to elevated concentrations of manganese (Mn) and iron (Fe). Such high HI values significantly exceed the acceptable risk threshold (HI = 1) and warrant urgent attention. Similar studies conducted in groundwater-dependent areas of Bangladesh have also reported elevated HI values among children. For example, Tajwar et al. (2024) observed HI values ranging from 1.2 to 21 for children in parts of the Satkhira, Khulna, and Bagerhat districts due to high Mn levels [67], while Rushdi et. al. (2023) reported HI values reaching up to 7.9 in arsenic- and manganese-affected regions of Moulvibazar [68]. The elevated HI in Mymensingh is thus consistent with trends observed in other floodplain aquifer systems in Bangladesh, where natural geochemical enrichment and inadequate water treatment contribute to chronic exposure risks [69], [70]. Additionally, children’s vulnerability is heightened by their lower body weight and higher water intake per body mass, leading to disproportionate exposure compared to adults [71]. These findings underscore the critical need for targeted risk mitigation, especially for children, through household-level treatment, community awareness, and water source prioritization.

In this study, health risk calculations were limited to the oral ingestion pathway, which is considered the primary exposure route for contaminants in drinking water. However, other exposure routes such as dermal absorption during bathing and inhalation of water aerosols during showering may also contribute to total exposure, particularly in settings where water is used frequently for personal hygiene. Although manganese (Mn) is not highly volatile, aerosolized particles and steam during hot water use can lead to inhalation exposure, which is increasingly recognized as relevant for neurotoxic metals like Mn [72]. Additionally, dermal absorption, though generally less significant for Mn and Fe due to their low skin permeability, may still be a concern for sensitive populations, especially with prolonged exposure or damaged skin [73].

Emerging evidence suggests that chronic low-level exposure to Mn via inhalation can have neurological effects, particularly in children, such as reduced IQ, behavioral issues, and motor skill impairment [74]. These findings underscore the need for a more holistic risk assessment framework that includes multiple exposure pathways. Future work should incorporate dermal and inhalation risk models (e.g., using USEPA’s Risk Assessment Guidance for Superfund - RAGS Part E), and assess household practices such as bathing frequency, water heating methods, and ventilation, which influence these exposures.

## 4. Conclusion

This study provides a comprehensive assessment of the present water supply status, drinking habits, and physicochemical quality of tap water. The findings indicate that tap water serves as the primary water source for the majority of respondents (89%), with hand tubewells catering to the remaining population. Despite a significant reliance on tap water, concerns regarding water quality and safety remain prevalent, as evidenced by 78% of respondents expressing worry about contamination. The physicochemical analysis of tap water samples reveals that while parameters such as pH, EC, TDS, and salinity remain within permissible limits, elevated concentrations of iron (Fe) and manganese (Mn) exceed national and international standards in a considerable number of samples. The high levels of these contaminants raise concerns regarding potential health risks, including gastrointestinal issues and neurotoxicity. Furthermore, the presence of total coliform bacteria in all sampled locations highlights microbial contamination, posing additional risks to public health. The study underscores the necessity of improved water treatment practices, as inconsistent or inadequate treatment was reported by a significant portion of respondents. Addressing water quality issues requires systematic monitoring, proper maintenance of supply infrastructure, and increased public awareness of water purification methods. Policymakers and relevant authorities should prioritize measures to enhance water quality, including upgrading supply systems, enforcing stricter water safety regulations, and promoting community-based interventions. Overall, while tap water remains a crucial resource for domestic use, ensuring its safety necessitates continuous assessment, investment in infrastructure, and public engagement. Future studies should explore seasonal variations in water quality and investigate the long-term health implications of contaminants detected in this study.

## Data Availability

All data produced in the present study are available upon reasonable request to the authors

## Data availability statement

Data will be made available on request.

